# *COCOH*: A Multimodal Deep Learning Framework for Cancer Risk Assessment of Oral Potentially Malignant Disorders

**DOI:** 10.64898/2026.01.04.26343399

**Authors:** John Adeoye, Yu-Xiong Su

**Affiliations:** AI for Head and Neck Oncology (AI4HNO) Group, University of Hong Kong, Hong Kong SAR, China; Division of Applied Oral Sciences and Community Dental Care, Faculty of Dentistry, University of Hong Kong, Hong Kong SAR, China; Division of Oral and Maxillofacial Surgery, Faculty of Dentistry, University of Hong Kong, Hong Kong SAR, China

**Keywords:** artificial intelligence, clinical decision-making, malignant transformation, precancerous conditions, oral cancer

## Abstract

Oral potentially malignant disorders (OPMD) are mucosal diseases with an increased risk of progression to cancer, although not all cases develop cancer in patients’ lifetimes. Although epithelial dysplasia (ED) grading is the current approach for assessing the risk of malignant transformation (MT), cancer risk prediction is limited by its subjective interpretation and inaccuracy. To address these challenges, this study developed ***COCOH*** (**<u>C</u>**omprehensive **<u>O</u>**ral **<u>C</u>**ancer predictor in **<u>O</u>**PMD using **<u>H</u>**istopathology and immunohistochemistry), a multimodal attention-based deep multiple instance learning (MIL) framework that integrates disease information, hematoxylin and eosin (H&E), and Keratin-13 (KRT13) immunohistochemistry whole-slide images (WSIs) to predict MT risk in OPMD. Trained on 2,780 WSIs and over 2.7 million image patches from cases with oral leukoplakia and oral lichenoid disorders, *COCOH* achieved an AUC of 0.904 (0.843-0.965) and Brier score of 0.089 (0.044-0.134) during testing, with excellent discrimination and calibration maintained at prospective validation (AUC: 0.951 [0.897-1], Brier score: 0.062 [0.002-0.122]). Notably, the deep learning model outperformed two ED grading systems in assessing cancer risk in oral leukoplakia, yielding a significant net reclassification improvement of 20.2%. Decision curve analysis confirmed *COCOH*’s clinical utility in identifying high-risk OPMD cases for intervention and close monitoring. These findings demonstrate that *COCOH* provides accurate and well-calibrated risk predictions for MT in OPMD, and its integration into clinical workflows could enhance prevention strategies and promote earlier detection of oral cancer.

## Introduction

Oral cancer is the most common malignancy of the head and neck, with over 390,000 cases recorded annually ^1^. Approximately one in two patients with oral cancer do not survive beyond five years due to factors such as comorbidities, diagnostic delay, and advanced tumor stage at presentation, which contribute to significant morbidity and mortality ^2,3^. Despite advances in cancer therapeutics, early detection and prevention remain the most effective strategies to reduce disease burden ^3^. Primary prevention measures that aim to reduce risk habit practices such as tobacco use, areca nut chewing, and heavy alcohol consumption have, increasingly been adopted in many regions ^4,5^. However, precancerous lesions known as oral potentially malignant disorders (OPMD) can also occur in individuals without these habits, making their detection and management critical for effective oral cancer prevention and early detection.

OPMD are diseases with an inherent predisposition for progression to oral cancer ^6^. While different subtypes exist, each with its own malignant potential, oral leukoplakia (OL) and oral lichenoid diseases (OLD – including lichen planus and lichenoid lesions) are the most common ^6,7^. Given that not all patients with OPMD develop malignancy, cancer-preventive therapies are not universally recommended for patients ^7^. Furthermore, heterogeneity in reported malignant transformation (MT) proportions (1.1 to 40.8% for OL and 0.44 to 3.2% for OLD) limits their use in guiding treatment decisions ^8,9^. This variability underscores the need for individualized cancer risk assessment to optimize clinical management for patients with OPMD.

Detecting and grading oral dysplasia following histopathology remains the most widely used approach to estimating malignant progression in OPMD ^10^. While effective at the population level, this method is unidimensional and often suboptimal for personalized risk assessment of patients with OL and OLD. Notably, a substantial proportion of patients without dysplasia (deemed low risk by current criteria) still develop oral cancer ^11^. Moreover, dysplasia grading suffers from limited accuracy/reproducibility, and a lack of consensus on grading conventions exist ^12^. It is also uncertain whether precursor cell categories or dysplasia grades are more predictive of cancer progression in clinical practice ^13^.

Assessing the risk of cancer development in OPMD is a complex clinical task that is often inferred indirectly from dysplasia grading, which is unreliable due to interobserver variability ^14,15^. Deep learning (DL) methods have been investigated for standardizing oral dysplasia assessment and discovery of computational features predictive of cancer risk from conventional hematoxylin and eosin (H&E) whole-slide images (WSI) ^16–20^. However, existing models often depend on manual (epithelial) annotations provided during preprocessing, rely on patch-level supervision with hard labels that can lead to false predictions due to noisy regions, lack contextual awareness and multiscale feature integration, and demonstrate suboptimal accuracy, precision, and calibration. Of note, these frameworks lack complementary patient-level clinical data or immunohistochemical markers to enhance cancer risk assessment, especially for patients with similar dysplasia grades. Keratin-13 (KRT13) has emerged as an important immunohistochemical indicator for comprehensive assessment of dysplasia and cancer risk in OL and OLD. Reduced expression of KRT13 is associated with an increased risk of cancer development and poorer progression-free survival ^21–23^. Therefore, in this study, we developed and validated a multimodal, attention-based deep multiple instance learning (MIL) system termed “<u>C</u>omprehensive <u>O</u>ral <u>C</u>ancer predictor in <u>O</u>PMD using <u>H</u>istopathology and immunohistochemistry (*COCOH*)” that integrates disease information, H&E WSIs, and immunohistochemistry (IHC) WSIs based on KRT13 for personalized, patient-level cancer risk assessments in OL and OLD.

## Methods

### Data sources

This multicenter study used 2,656 OL and OLD tissue slides from 949 patients across 2 centers to predict the risk of cancer development in OPMD. Cohort included those with OL and OLD who were managed at the Queen Mary Hospital (QMH) and the Prince Philip Dental Hospital (PPDH) in Hong Kong from January 2003 to December 2022. OLD comprised oral lichen planus and oral lichenoid lesions, and the criteria for patient selection were based on the WHO 2020 definition ^6^. Patients with other OPMD or oral malignancies at the time of diagnosis or those without initial biopsies or archival tissues were excluded from this study. For all patients, definitive diagnoses (OL or OLD) and clinical outcomes (cancer development or otherwise as of November 18, 2025) were extracted from the electronic medical records of both institutions. MT was defined as biopsy-proven oral squamous cell carcinoma at the same anatomic site as OPMD after six months of index biopsy. Those who did not develop cancer had at least 34 months of follow-up. All formalin-fixed paraffin-embedded (FFPE) tissues from surgical biopsies performed before cancer development or the last follow-up were obtained. Characteristics of the study cohort have been detailed in a previous study ^21^. Overall, 534 OL cases (781 tissue samples) and 415 OLD cases (547 tissue samples) with MT rates of 12.4% and 9.9% were included (mean follow-up of 97.4 months). The MT rate for OLD may be elevated in this study due to patient selection according to FFPE availability, which may enrich for lesions of clinical concern. Ethics approval was obtained from the Institutional Review Board of the University of Hong Kong/Hospital Authority Hong Kong West Cluster (Reference no: UW 22-789), and informed consent was waived due to the retrospective nature of this study.

### H&E staining, immunostaining, and whole-slide imaging

Serial 5µm FFPE sections of OL and OLD were processed for H&E staining and KRT13 immunostaining. H&E staining procedures included deparaffinization with xylene (twice) and graded alcohol (100% (twice), 95%, and 70%), staining with hematoxylin for 3 minutes, differentiation using 0.1% HCl in 70% ethanol for 3 seconds, bluing in 1% ammonia for 1 minute, and additional staining with Eosin Y for 40 seconds. Slides were then air-dried and mounted with coverslips. KRT13 immunostaining procedures have been documented in a previous report ^21^ and detailed in **Supplementary Methods**. H&E (n=1328) and KRT13 (n=1328) sections were digitized using brightfield settings of the high-throughput PhenoImager HT system (Akoya Biosciences, MA, USA) at 40× magnification (0.25µm/pixel). Other resolutions i.e., 20×, 10×, and 5×, were 0.5 µm/pixel, 1 µm/pixel, and 2 µm/pixel. Overall, 2,656 WSIs were scanned and used for *COCOH*’s development and validation.

### Data preprocessing

We performed label encoding for disease information and MT status in spreadsheets. H&E and KRT13 WSIs were anonymized and matched to annotated patient outcomes and disease information (OL or OLD) recorded in a spreadsheet. Otsu’s thresholding was performed to delineate tissue areas and remove background. Nonoverlapping patches of 256 × 256 pixels were extracted from tissue areas, and those containing <90% white background were retained. Multiscale tiling (5×, 10×, and 20×) was performed for H&E WSIs to ensure context awareness and representation of architectural and cytologic features, while mimicking pathologists’ workflow when reviewing histopathology slides of patients with OPMD ^24^. Stain normalization was not performed to mitigate batch- and site-specific factors in model performance ^25^. For KRT13 WSIs, we only performed single-scale tiling at 5× because the protein is an epithelial differentiation marker whose reduced expression (associated with cancer progression) would be apparent at a low magnification ^21,22^. Images were resampled offline using a stratified 75:15:10 train-validation-test split at the patient level to prevent data leakage. Patients were first stratified by OPMD subtype and further divided within each type by their cancer progression status. Afterward, the splitting proportion was applied to ensure that the prevalence of cancer development in OPMD remained for the disease subtypes.

### *COCOH*’s architecture

*COCOH* is a multimodal deep learning framework that integrates disease information, histopathological features, and KRT13 features for cancer risk assessment in OPMD. *COCOH*’s framework uses a hybrid supervised learning approach where OPMD subtypes are processed using fully supervised learning, while histopathological and IHC features are represented using attention-based weakly supervised learning. Here, WSIs are viewed as bags of features extracted from H&E/KRT13 IHC image patches, which are aggregated independently and modeled to outcomes available at the WSI level. Furthermore, to enhance cancer risk assessment, the model leverages a two-stage multimodal fusion strategy. Histopathological and IHC features are first integrated via intermediate fusion, after which the combined WSI representations are fused with the disease identifier before being passed to a classifier to estimate the MT probability of the OPMD. Overview of the DL architecture is in **Fig. 1**.

**Figure 1:**
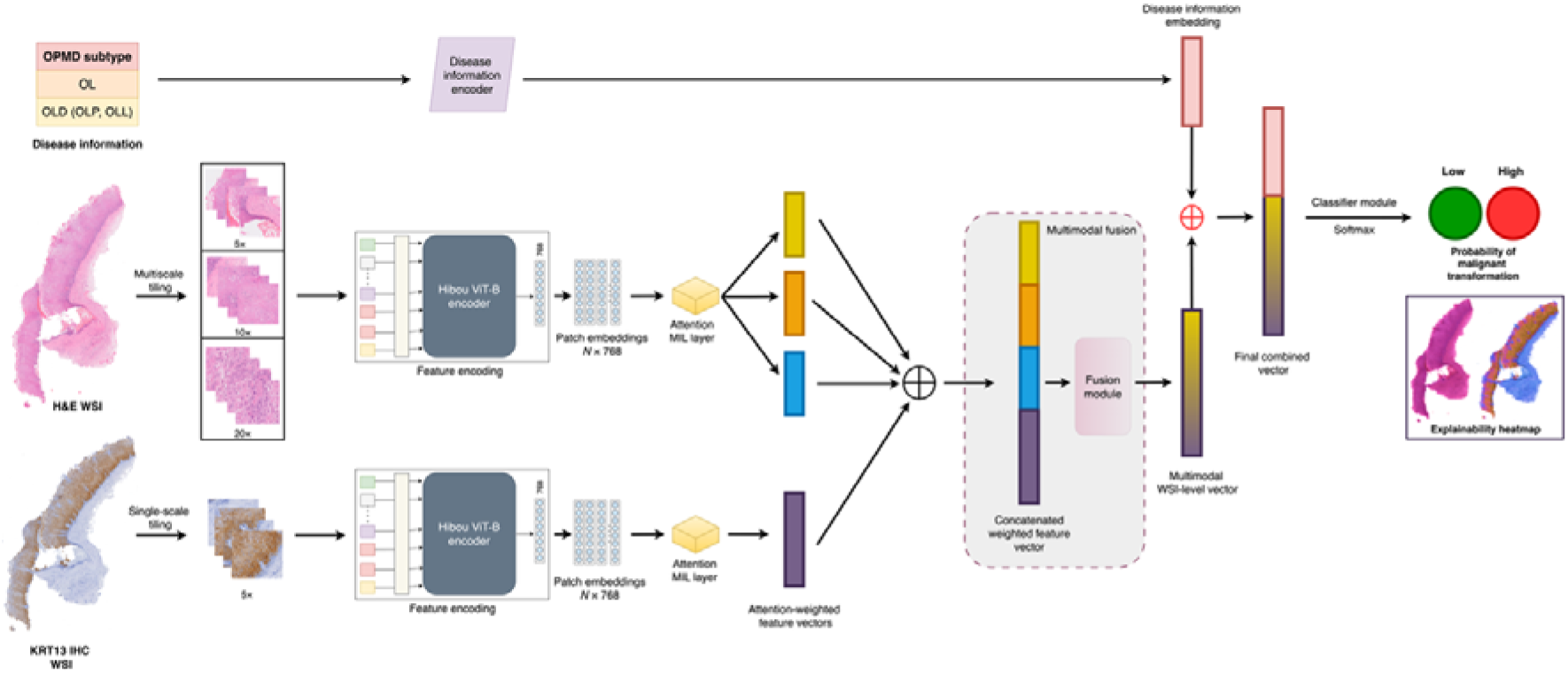
Overview of *COCOH*’s architecture. The model takes as input the clinical subtype of OPMD and digitized hematoxylin and eosin (H&E) and KRT13 immunohistochemistry (IHC) whole-slide images (WSIs). Disease information is processed through a fully connected layer to generate a disease embedding. H&E WSIs are tiled at multiple resolutions (5×, 10×, and 20×), while KRT13 WSIs are tiled at 5×. Hibou-B serves as the feature encoder, and patch embeddings from each scale or modality are aggregated using attention-based multiple-instance learning. Attention-weighted vectors are concatenated and passed through a fusion module to produce a multimodal WSI-level vector, which is then fused with the disease embedding. The final combined vector is fed into a classifier head to estimate the probability of malignant transformation. Explainability heatmap is generated using patch-level attention scores for each modality.

#### Feature encoding

To generate informative feature representations from H&E and KRT13 IHC image tiles, we used Hibou-B ^26^, a pathology foundational model based on the Vision Transformer (base variant) architecture. The model was pretrained with the DINOv2 self-supervised learning method on >1 million diverse WSIs. We selected Hibou because its pretraining dataset included both H&E and non-H&E WSIs, with a large proportion of WSIs from head and neck tissues compared to other publicly available models. During feature extraction, H&E/KRT13 IHC image patch were resized and normalized. Hibou was then run in evaluation mode to convert preprocessed tiles to a feature embedding of 768 dimensions obtained from the [CLS] token of the final transformer layer. This embedding serves as a global representation of each WSI patch. This process was then repeated for all patches from a given WSI (W), and the resulting feature vectors were stacked into an N × 768 feature matrix (bags of features), where N is the total number of image patches obtained from W.

#### Attention-based multiple instance learning

For H&E and KRT13 modalities, we used tanh-sigmoid gated attention MIL pooling inspired by Ilse et al ^27^ to aggregate patch-level features for predicting MT probability at the slide level. This trainable method uses two parallel neural networks with hyperbolic tangent and sigmoid activation functions that compute an intermediate attention vector and a gate vector of the same dimension, which are multiplied elementwise. Gated attention vector is then passed through a dense layer to compute raw scalar attention scores for each instance (feature embedding) in the WSI, which are softmax-normalized and used to weigh/scale patch feature embeddings based on their importance to OPMD cancer risk. Weighted feature embeddings are then summed to obtain WSI-level representations for H&E and IHC modalities.

Attention scores can be represented mathematically as:

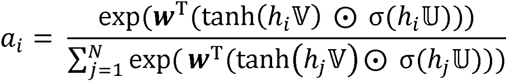

Given:

*N* = number of instances per bag

*D* = hidden dimension of instance features

*L* = hidden dimension of the attention mechanism

*H* = [*h*_1_, *h*_2_, *h*_3_, …, *h_i_*] ϵ ℝ^*N×D*^ are *N* instance features (for each WSI) projected to *D*.

Let:

Tangent transformation (τ) for instance (*h_i_*) = tanh(*h_i_*𝕍)∈ℝ^*L*^, where 𝕍 ∈ ℝ^*D×L*^ is the learnable weight matrix for applying tanh non-linearity.

Sigmoid transformation (Σ) for *h_i_* = σ(*h_i_*𝕌)∈ℝ*^L^*, where 𝕌∈ ℝ*^D×L^* is the learnable weight matrix for applying sigmoid non-linearity.

Then:

*G_i_* = τ_*i*_ ⊙ Σ_*i*_, where *G_i_* is the gated vector and ⊙ denotes element-wise multiplication

Raw attention scores, α*^r^_i_* = ***w**^T^G_i_*, where ***w***∈ℝ^*L*×1^ is a learnable parameter for computing scalar attention weights for instance.

Normalized attention scores, 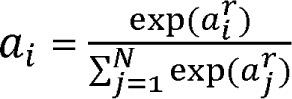

To obtain WSI-level representation, given attention scores (α) and feature embeddings *H*, MIL aggregation per WSI modality is:

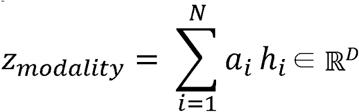

#### Two-stage multimodal fusion

*COCOH*’s architecture includes a two-stage fusion strategy to integrate the two WSI modalities and disease information. First, WSI-level representations per modality (*z_he_*, *z*_*krt*13_∈ℝ^*D*^) are concatenated. Given the multiscale aggregated embeddings for the H&E modality (5×, 10×, 20×), the concatenated WSI-modality representation is represented as:

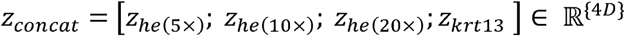

Concatenated representation is then fused via a dense layer with ReLU activation to give a single multimodal WSI-level vector that integrates multiscale H&E and KRT13 modalities which is:

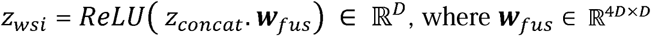

The scalar disease information (*d* ϵ ℝ^1^), which is important for contextualizing fused H&E and KRT13 WSI modalities, is passed through a fully connected layer with ReLU to output an embedding (*z_disease_*) projected to *D*.

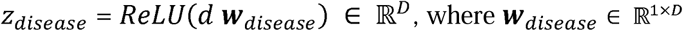

The multimodal WSI-level vector is then concatenated with the disease information embedding as *z_combined_* = [*z_wsi_*; *z_disease_*] ϵ ℝ^2*D*^ and this final combined vector is passed to a classifier head to provide logits that are converted to MT probabilities using softmax.

### Model training and cross-validation

To ensure *COCOH*’s stability and selection of optimal hyperparameters, five-fold cross-validation (CV) was performed with the training dataset (4 parts training, 1 part validation) while monitoring the loss on the validation partition. List of hyperparameters tuned is in **Table S1**. Training was achieved with a hidden dimension of 256, batch size of 1, Adam with decoupled weight decay optimizer (AdamW, weight decay=0.00001), maximum epoch of 100, learning rate of 0.0001, dropout of 0.25 and focal loss (modified cross-entropy) to handle imbalance in the outcome data. For the loss function, we set a (balancing) and y (focusing) hyperparameters at 0.75 and 2.0. Early stopping was implemented after a minimum epoch of 5 if no decrease in validation loss is observed after 10 epochs. Mean and standard deviation (SD) of the area under the receiver operating characteristic curve (AUC), area under the precision-recall curve (AUPRC), and Brier scores were calculated across the folds to determine training performance and stability.

### Model performance analysis

To evaluate *COCOH*’s performance, we retrained the model on the entire training dataset using the same parameters as CV.

#### Internal validation, ablation experiments, and alternate architectures

Discrimination and calibration were assessed on the internal validation dataset using AUC and Brier score. To evaluate the specific contribution of multimodal fusion, we performed ablation experiments by removing different modalities from *COCOH* and determined the AUCs and Brier scores of the resulting model. This involved setting z_modality_ to zero in the training and validation datasets. Matched H&E and KRT13 WSIs were available for all patients, and modality dropout was not performed during training. However, since KRT13 WSIs may be unavailable in clinical settings compared to H&E, we implemented zero substitution of KRT13 input embeddings, while maintaining the same shape as the 5× H&E embedding to allow the model to generate a stable placeholder if this modality is absent. We then computed the AUC and Brier score of the model when all KRT13 WSIs of the validation dataset were dropped to quantify the impact on performance at internal validation. To ensure that the different components and strategies for developing *COCOH* are optimal, this study also benchmarked the model’s performance (AUC and Brier score) against other architectures trained and evaluated using the internal validation set. Details of these variant models are in **Supplementary methods**.

#### Model Testing

Generalizability assessment was done on the independent testing dataset. Performance metrics for all OPMD and separately for OL and OLD subtypes included AUC, AUPRC, and Brier score. 95% confidence intervals (CI) were also computed. Based on Youden’s index, we selected optimal thresholds to stratify cancer risk in OL and OLD using the internal validation dataset. These thresholds were then applied at testing to calculate the recall, specificity, precision, and negative predictive values (NPV) of *COCOH*. Potential algorithmic bias was assessed by comparing the AUC and AUPRC between OL cases with and without dysplasia.

#### Benchmarking against dysplasia grading in OL

We benchmarked COCOH’s test performance against WHO and binary dysplasia grading systems using similar metrics (AUC, recall, specificity, precision, and negative predictive value). Net reclassification improvement (NRI) of *COCOH* was also determined compared to binary grading for OL cases. Mathematical representations of NRI are in **Supplementary Methods**.

#### Prospective validation

Additional evaluation of the model was performed using an independent cohort of consecutive OPMD cases (n=124) treated at PPDH between January 1 and December 31, 2023, that were followed prospectively. This cohort had an average follow-up of 27.3 months. H&E staining, KRT13 IHC, and whole-slide imaging were conducted using protocols similar to those of the development cohort (18 months after). We also determined the discrimination and calibration performance of *COCOH* using similar metrics above.

### Net benefit and Explainability analysis

Net benefit was determined separately for OL and OLD using decision curve analysis of predicted probabilities obtained for the testing and prospective validation cohorts. All possible threshold probabilities were explored, and all values were used to plot decision curves for *COCOH*, which were compared to reference decision curves. Explainability was implemented at 5× and 20× scales of the H&E modality and 5× scale of the KRT13 modality using intrinsic attention weights computed for each patch. For each WSI, patch-level attention scores were obtained from the trained model, normalized using percentile scaling, and interpolated to the original resolution to generate a probability heatmap for visualizing patches that contributed to the predicted probabilities. Patch probability heatmaps were then superimposed on WSIs per modality. WSIs of 10 random patients with correct predictions at testing (5 OL and 5 OLD, 60% event rate) were reviewed by two pathologists to identify histologic and immunohistochemical features associated with high/low MT probabilities. Likewise, 7 patients (5 OL and 2 OLD, 42.9% event rate) in the same cohort whose predictions were incorrect were reviewed to identify histologic and immunohistochemical features in the WSIs contributing to misclassification. To confirm the importance of patch-level features, we removed the vectors with the top ten attention scores per modality for the test and prospective validation data before recalculating the AUC and Brier scores to ensure these areas accurately informed predictive performance.

### Site-specific factors analysis

To evaluate the impact of whole-slide imaging systems on model performance, we randomly selected 124 tissue slides (62 corresponding H&E and KRT13) from the test and prospective validation sets and rescanned them at 20× on a Nikon Eclipse Ti2-E inverted microscope. WSIs were tiled into 256×256 patches. The 20× scans were downsampled by factors of two and four during tiling to yield 10× and 5× patches. Patches were encoded using Hibou-B and processed via *COCOH*’s attention MIL pooling, modality fusion, and prediction module. Performance on these rescanned WSIs was assessed using AUC and Brier score and compared with WSIs of the same slides (matched by slide ID) acquired with the Akoya PhenoImager HT system. Differences in the AUC of the model between the imaging modalities were assessed using DeLong’s test, with p-values below 0.05 considered as statistically significant.

## Results

### Model stability and internal validation

*COCOH* was trained using 1,926,952 WSI patches (1,822,514 multiscale H&E, 104,438 KRT13) (**Table S2**). The model demonstrated strong performance for predicting MT in OPMD at CV, with a mean (SD) AUC of 0.881 (0.030), AUPRC of 0.667 (0.057), and Brier score of 0.092 (0.011) (**Fig. 2A-C**). Across clinical subtypes, performance was slightly higher for OL than for OLD, with mean AUCs of 0.881 (0.044) and 0.878 (0.007). Similarly, mean AUPRC was higher for OL than OLD (0.709 [0.068] vs. 0.582 [0.038]). In contrast, model calibration was better for OLD (Brier score: 0.076 [0.013] vs 0.103 [0.011] for OL). Overall, standard deviation values following five-fold CV indicated that OLD predictions were slightly more stable than OL predictions.

**Figure 2:**
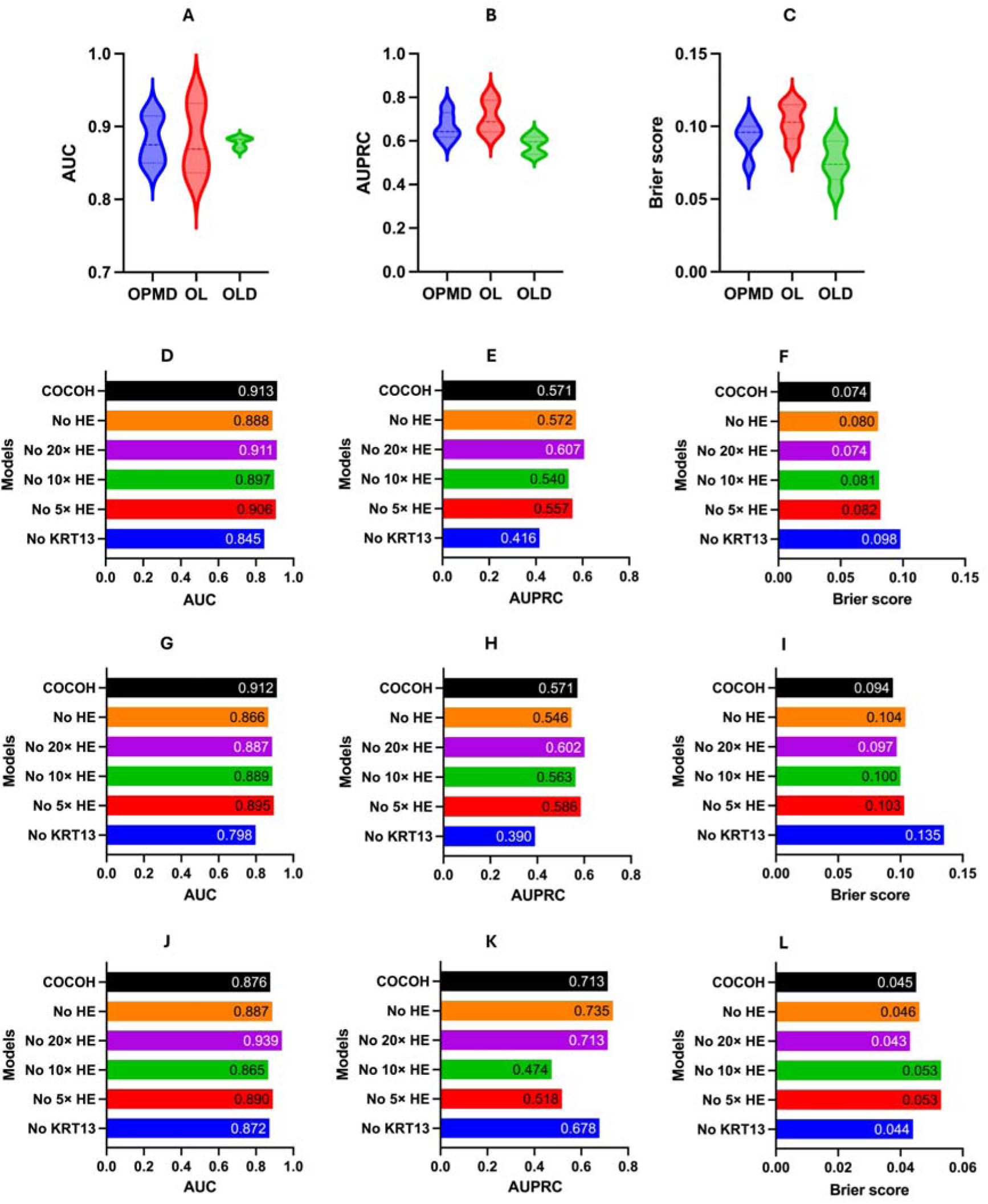
Five-fold cross-validation (CV) and internal validation performance of *COCOH* vs unimodal and variant models. (A-C) Violin plot showing *COCOH*’s (A) AUC, (B) AUPRC, and (C) Brier score in predicting cancer risk of OPMD, OL, and OLD across five CV folds. (D) Bar plot comparing *COCOH*’s **AUC** with those of five unimodal and variant models in predicting cancer risk in **OPMD** at internal validation. (E) Bar plot comparing *COCOH*’s **AUPRC** with those of five unimodal and variant models in predicting cancer risk in **OPMD** at internal validation. (F) Bar plot comparing *COCOH*’s **Brier score** with those of five unimodal and variant models in predicting cancer risk in **OPMD** at internal validation. (G) Bar plot comparing *COCOH*’s **AUC** with those of five unimodal and variant models in predicting cancer risk in **OL** at internal validation. (H) Bar plot comparing *COCOH*’s **AUPRC** with those of five unimodal and variant models in predicting cancer risk in **OL** at internal validation. (I) Bar plot comparing *COCOH*’s **Brier score** with those of five unimodal and variant models in predicting cancer risk in **OL** at internal validation (J) Bar plot comparing *COCOH*’s **AUC** with those of five unimodal and variant models in predicting cancer risk in **OLD** at internal validation (K) Bar plot comparing *COCOH*’s **AUPRC** with those of five unimodal and variant models in predicting cancer risk in **OLD** at internal validation (L) Bar plot comparing *COCOH*’s **Brier score** with those of five unimodal and variant models in predicting cancer risk in **OLD** at internal validation.

Internal validation performance is presented in **Figs. 2D-L**. *COCOH* had an AUC (95% CI) of 0.913 (0.873-0.953), an AUPRC (95%CI) of 0.571 (0.501-0.641), and a Brier score (95%CI) of 0.074 (0.037-0.111) for OPMD (**Fig. 2D-F**). OPMD subtype analysis showed that the model differentiates high and low-risk patients better for OL compared to OLD (AUC: 0.912 [0.86-0.964] vs 0.876 [0.804-0.948]) (p-value: 0.7015; **Figs. 2G, J**). Similar to CV performance, calibration was higher in OLD than OL, which may be due to lower MT prevalence in OLD (**Figs. 2I, L**). Interestingly, AUPRC was higher for OLD (0.713 [0.614-0.812]) than OL (0.571 [0.480-0.660], likely reflecting differences in MT prevalence between the CV and validation cohorts. On the internal validation set, 25% and 30% were defined as optimal thresholds for cancer risk stratification in OL and OLD, respectively.

Detailed comparison of alternative approaches performed during model development on the internal validation set is presented in **Supplementary Results** and **Fig S1**.

### Unimodal baseline comparisons

Ablation experiments revealed that KRT13 contributed most to *COCOH*’s performance. Removing KRT13 features resulted in the largest decline, with AUC decreasing by 0.068 (95% CI: -0.104 to -0.032), AUPRC by 0.155 (95% CI: -0.206 to -0.104), while Brier score increased by 0.024 (95% CI: 0.002-0.046) across all OPMD cases (**Fig. 2D-F**). Stratified analysis revealed that this performance drop was more marked for MT predictions in OL than in OLD (**Figs. 2G-L)**.

Removing H&E features at different resolutions or altogether had variable effects on model performance (**Figs. 2D-L**). Excluding all H&E features or only the 20× scale reduced model performance for OL only, while removing 5× or 10× H&E features decreased *COCOH*’s performance for OL and OLD. Overall, these variant models showed imbalanced performance for risk assessment of both disorders. None of these unimodal or bimodal variants outperformed the full *COCOH* model, which was then retained for further analysis.

### Missing modality handling

This study also evaluated the zeroing vector method used to learn missing KRT13 features in *COCOH*. When KRT13 vectors in the validation dataset were replaced with null vectors, the AUC and AUPRC dropped by -0.052 (95% CI: -0.083 to -0.021) and -0.151 (95% CI: -0.202 to -0.100) respectively for all OPMD. These reductions in performance were smaller than when the KRT13 vectors were completely removed (zero modality weights), indicating that this approach preserved discrimination. However, this improvement resulted in poorer calibration (**Fig. S2A-C**). Subtype analysis also confirmed improvements in AUC and AUPRC for OL and OLD when zero vectors were used in learning the missing KRT13 modality (**Figs S2 D-I**).

### Overall performance of *COCOH*

*COCOH* exhibited outstanding performance at independent testing, achieving an AUC of 0.904 (0.843-0.965), an AUPRC of 0.799 (0.736-0.862) and a Brier score of 0.089 (0.044-0.134) for all OPMD cases at independent testing (**Figs. 3A-G**). Discriminative performance was similar for OL (AUC: 0.897 [0.829-0.966]) and OLD (AUC: 0.9 [0.736-1]) (p-value: 0.9685; **Figs. 3B, C**). However, a higher AUPRC was obtained for OL than OLD (0.821 [0.745-0.897] vs. 0.745 [0.632-0.858]) (**Figs. 3F, G**). MT probabilities were better calibrated for OLD compared to OL (Brier: 0.047 [0-0.102] vs. 0.1133 [0.031-0.196] for OL).

**Figure 3:**
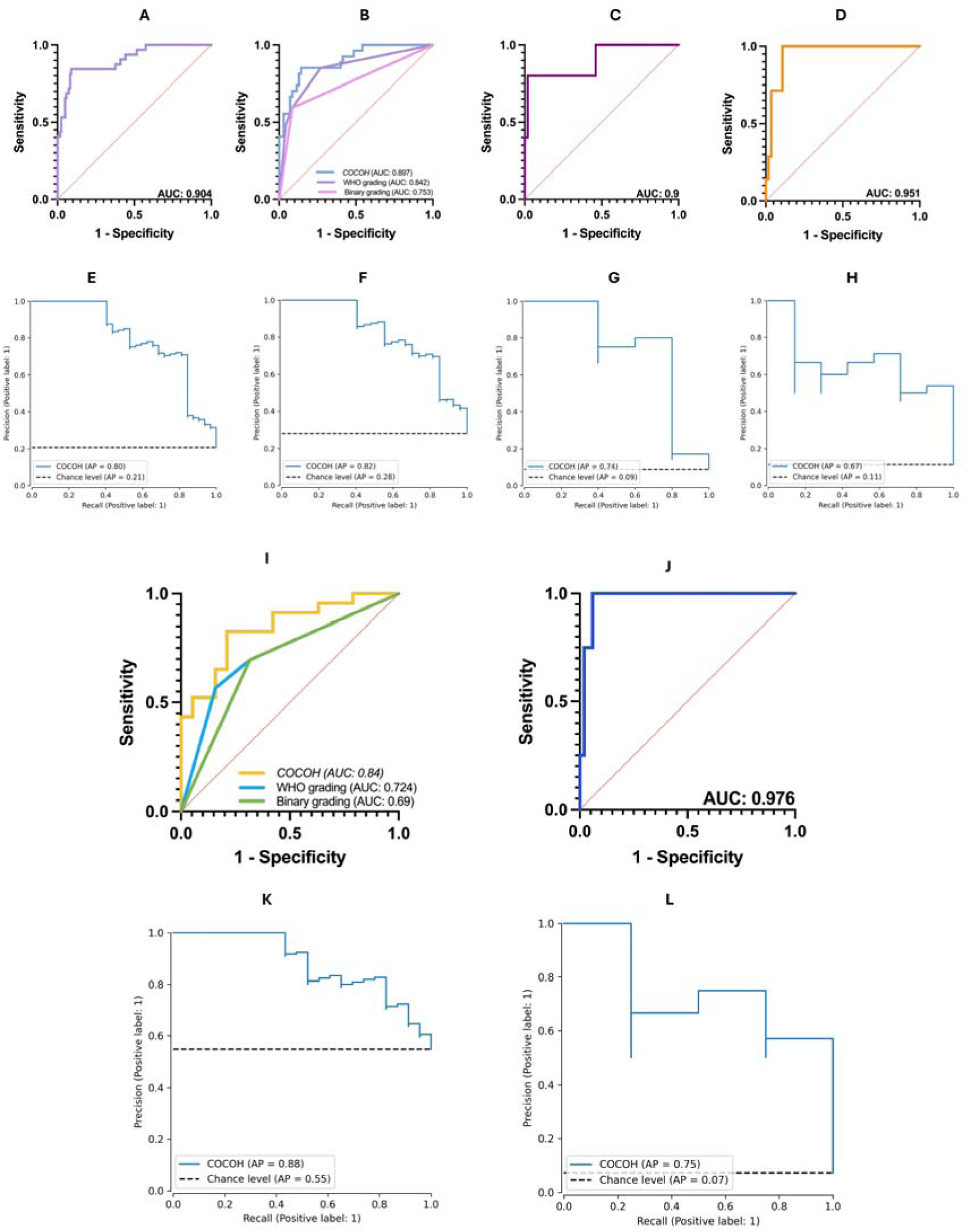
Testing and prospective validation performance of *COCOH*. (A) **ROC curve** indicating *COCOH*’s discriminative performance on all **OPMD** cases at **testing** (AUC: 0.904). (B) **ROC curves** comparing the discriminative performance of *COCOH* compared to WHO and binary dysplasia grading system in predicting cancer risk in **OL** at **testing**. *COCOH* achieved better AUC of 0.897 compared to AUCs of 0.842 and 0.753 obtained for WHO and binary grading. (C) **ROC curve** indicating *COCOH*’s discriminative performance on **OLD** cases at **testing** (*AUC: 0.90*). (D) **ROC curve** indicating *COCOH*’s discriminative performance on all **OPMD** cases at **prospective validation** (*AUC: 0.951*). (E) **Precision-recall curve** of *COCOH* on all **OPMD** cases at **testing** (F) **Precision-recall curve** of *COCOH* on **OL** cases at **testing** (G) **Precision-recall curve** of *COCO*H on **OLD** cases at **testing** (H) **Precision-recall curve** of *COCOH* on all **OPMD** cases at **prospective validation** (I) **ROC curves** comparing the discriminative performance of *COCOH*, WHO grading, and binary grading on **OL** cases with dysplasia at **testing**. *COCOH* achieved better AUC of 0.840 compared AUCs of 0.724 and 0.690 obtained for WHO grading and binary grading, respectively. (J) **ROC curves** comparing the discriminative performance of *COCOH* among **OL** cases without dysplasia at **testing** (*AUC: 0.976*). (K) **Precision-recall curve** of *COCOH* on **OL** cases with dysplasia at **testing**. (L) **Precision-recall curve** of *COCOH* on **OL** cases without dysplasia at **testing**.

Using the optimal probability threshold from internal validation, the recall, specificity, precision, negative predictive value, and balanced accuracy of *COCOH* for cancer risk stratification in OL were 85.2%, 85.7%, 69.7%, 93.8%, and 85.5%. Likewise, *COCOH*’s recall, specificity, precision, negative predictive value, and balanced accuracy for OLD were 80%, 98.1%, 80%, 98.1%, and 89% respectively.

At prospective validation, *COCOH* maintained excellent discrimination with an AUC of 0.951 (0.897-1), which was consistent with testing performance (**Fig. 3D**). AUPRC and Brier scores were 0.669 (0.552-0.782) and 0.062 (0.002-0.122), respectively (**Fig. 3H**). For cancer risk prediction in OL, the model achieved an AUC, Brier score, recall, specificity, and balanced accuracy of 0.925, 0.085, 100%, 80.6%, and 90.3%. Though none of the OLD cases in the prospective cohort developed MT, *COCOH* correctly predicted all as low risk, yielding 100% specificity and a Brier score of 0.009.

### *COCOH* vs. dysplasia grading for OL risk assessment

*COCOH* consistently outperformed WHO and binary dysplasia grading systems, which represent the current method of assessing cancer risk in OL. At independent testing, the DL model achieved an AUC that was 5.5% higher than WHO grading (95% CI: 1-10%) and 14.4% higher than binary grading (95% CI: 7.4-21.4) (**Fig. 3B**). Moreover, compared to binary grading, *COCOH* had significantly better recall (85.2 vs. 59.3%), NPV (93.8 vs. 85.3%), and balanced accuracy (85.5 vs. 75.4%).

NRI analysis further supported the findings above at independent testing. Ten cases misclassified as low risk by binary grading were correctly reclassified as high risk by *COCOH*, while three high-risk cases on binary grading were incorrectly downgraded by *COCOH*. Conversely, three cases misclassified as high risk by binary grading were correctly downgraded to low risk by *COCOH*, and seven correctly classified as low-risk cases by dysplasia grading were upgraded to high risk by *COCOH*. For OL cases with MT, *COCOH* significantly improved risk assessment by 25.9% (95% CI: 9.4-42.5%). For OL cases without MT, NRI was -5.7% (−14.4 to 0.03%), which was not statistically significant. The overall NRI was 20.2% (95% CI: 5.1-35.3%), indicating a general improvement in *COCOH*’s performance over binary grading.

### Algorithmic fairness and subgroup analysis in OL

*COCOH* showed strong discrimination for OL cases with or without dysplasia (**Figs. 3I, J**). However, its ability to predict cancer risk in OL without dysplasia was significantly better than for OL with dysplasia, with an AUC difference of 0.136 (95% CI: 0.068-0.204, p=0.0173). For both subgroups, AUPRCs were above the chance level (**Figs. 3K, L**).

Notably, *COCOH* outperformed WHO grading and binary dysplasia grading in predicting cancer risk of OL cases with dysplasia (AUC: 0.84 [0.721-0.959] vs. 0.724 [0.568-0.881] vs. 0.69 [0.526-0.854]) (**Fig. 3I**). For these dysplasia cases, *COCOH* had a higher recall, specificity, precision, NPV, and balanced accuracy of 82.6%, 79%, 82.6%, 79%, and 80.8% compared to binary dysplasia grading (recall: 69.6%, specificity: 68.4%, precision: 72.7%, NPV: 65%, balanced accuracy: 69%).

### Net benefit and explainability analysis

Decision curve analysis revealed that *COCOH* provides potential net clinical benefit if employed for cancer risk assessment of OPMD, supporting its use to identify cases that require intervention and close monitoring at testing and prospective validation (**Fig. 4**). Across all threshold probabilities, the decision curves for the DL model were consistently higher than those of reference curves for OL and OLD. Furthermore, *COCOH* showed superior net benefit at clinically relevant threshold probabilities above 15% compared to WHO and binary dysplasia grading in OL, indicating improved patient selection for treatment or follow-up.

**Figure 4:**
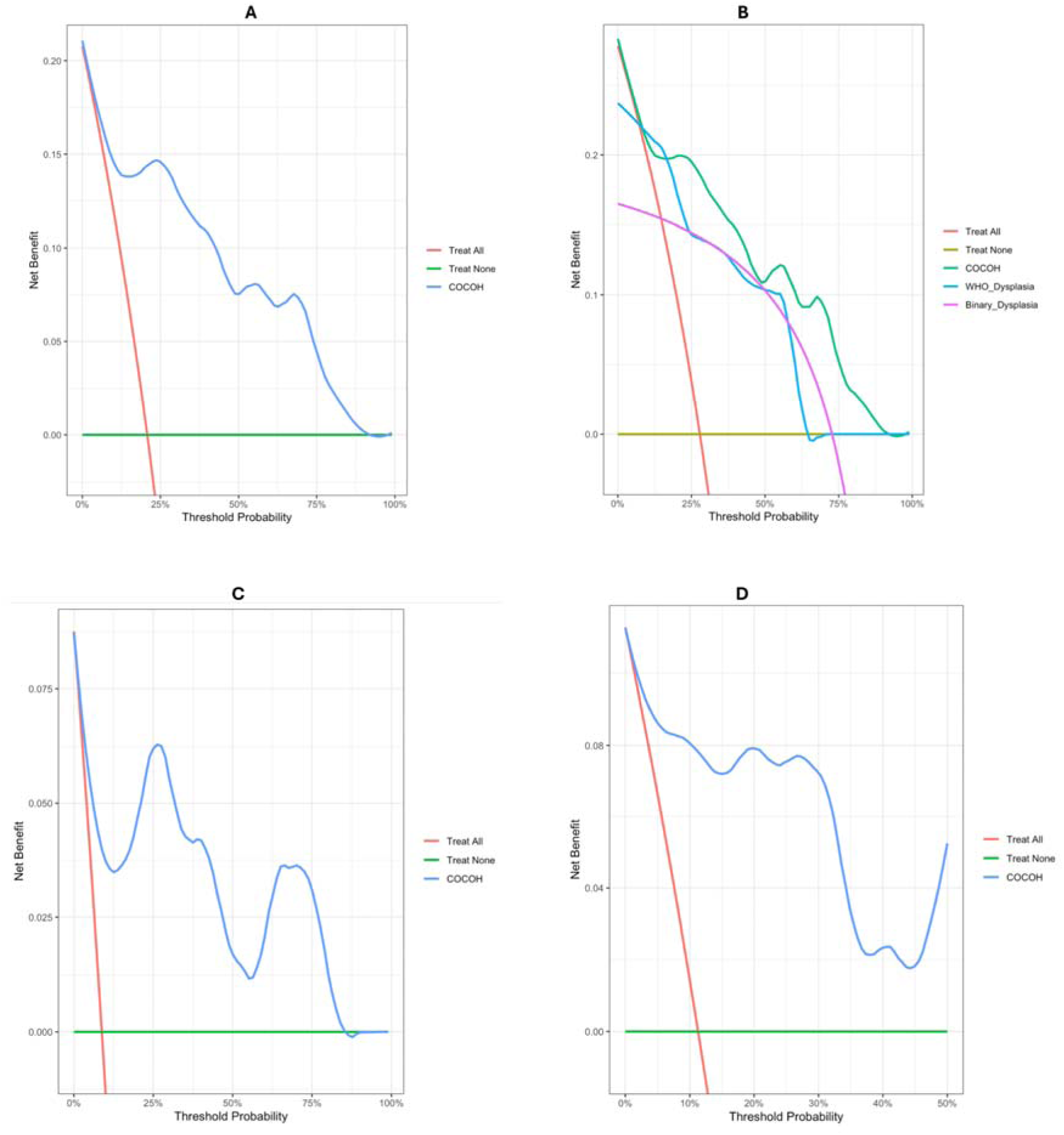
Net clinical benefit of *COCOH*. (A) Decision curves of *COCOH* compared to ‘treat all’ and ‘treat none’ reference decision curves for all **OPMD** cases at **testing**. *COCOH* had higher decision curves indicating superior potential clinical benefit if used to select OPMD cases for intervention and close monitoring. (B) Decision curves of *COCOH* compared to decision curves of WHO dysplasia grading and binary grading, and reference decision curves for **OL** cases at **testing**. (C) Decision curves of *COCOH* compared to reference decision curves for **OLD** cases at **testing**. (D) Decision curves of *COCOH* compared to reference decision curves for **OPMD** cases at **prospective validation**.

**Figure 5:**
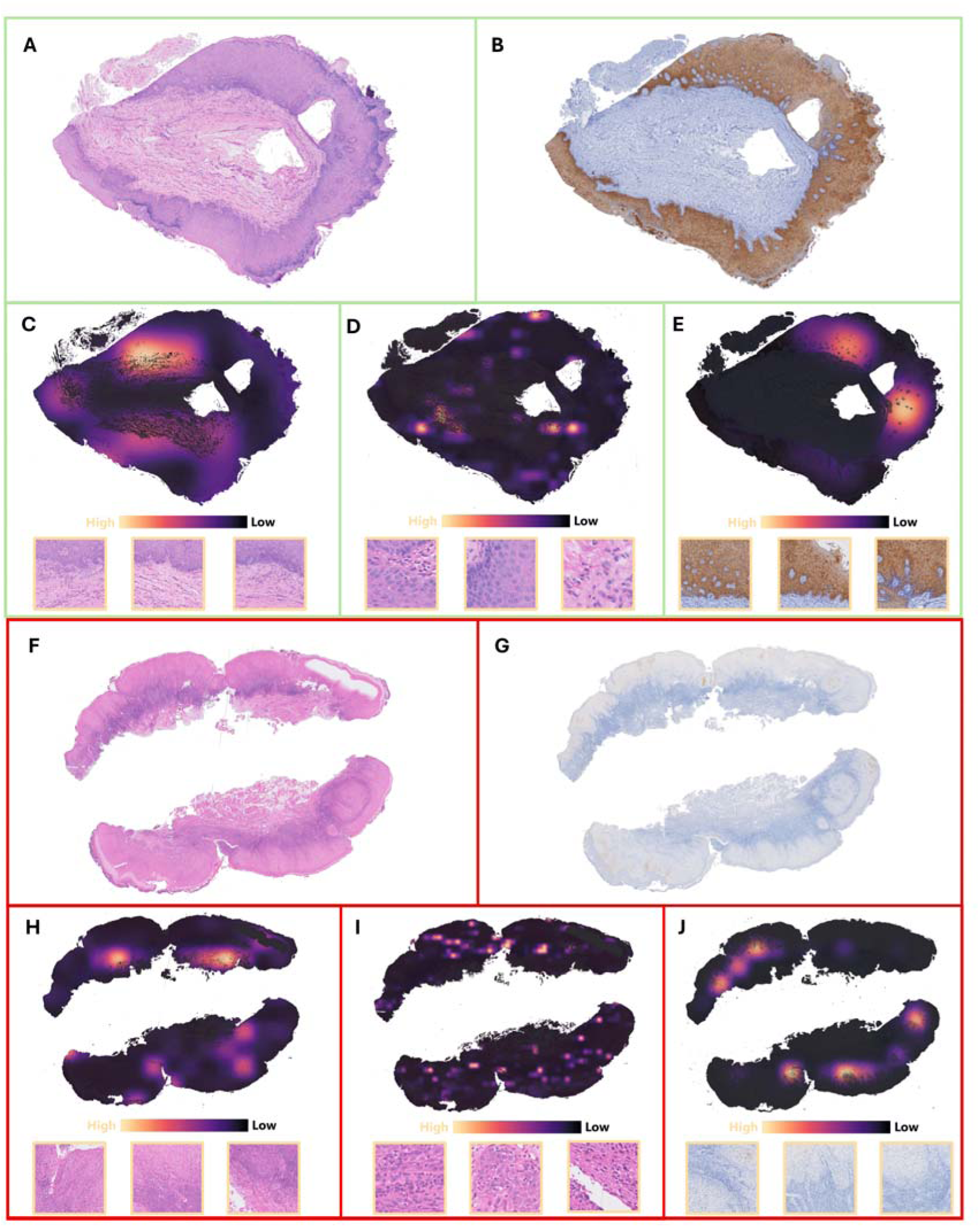
Multimodal explainability heatmaps based on *COCOH*’s normalized attention scores for WSIs with correct predictions. (A) H&E WSI for an **OL** case **without MT** after 39 months of follow-up. *COCOH* predicted an MT probability of 3%. (B) Corresponding KRT13 IHC WSI of patient in A (C) Attention heatmaps and top patches of H&E WSIs at 5× resolution showing important areas that influenced *COCOH*’s prediction. (D) Attention heatmaps and top patches of H&E WSIs at 20× resolution (E) Attention heatmaps and top patches of KRT13 WSIs at 5× resolution (F) H&E WSI for an **OLD** case **with MT** after 51 months follow-up. *COCOH* predicted an MT probability of 81% (G) Corresponding KRT13 IHC WSI of patient in F (H) Attention heatmaps and top patches of H&E WSIs at 5× resolution showing important areas that influenced *COCOH*’s prediction (I) Attention heatmaps and top patches of H&E WSIs at 20× resolution (J) Attention heatmaps and top patches of KRT13 WSIs at 5× resolution.

Multimodal explainability heatmaps based on *COCOH*’s attention scores for each modality for correct predictions and misclassification are shown in **Figs. S3 and 5**. Analysis of high-attention patches revealed that *COCOH* focuses more on the epithelium-connective tissue junction, irregular epithelial stratification, rete ridges, and keratinization. Notably, the attention maps for KRT13 WSIs also highlighted areas of aberrant staining intensity (increased and reduced) areas, which were associated with MT risk. When the patch feature vectors with the top ten attention scores were removed, the AUC decreased from 0.904 to 0.823 at testing and from 0.951 to 0.899 at prospective validation. Likewise, the Brier score at testing and prospective validation increased from 0.089 to 0.13 and from 0.062 to 0.085, supporting the validity of the explainability maps.

### Site-specific factor analysis

*COCOH*’s discrimination was unaffected by differences in WSI scanner type. When evaluated on slides scanned with a different system from that used for training, the model achieved an AUC of 0.870 (95% CI: 0.786-0.954), which is comparable to the AUC of 0.864 (95% CI: 0.779-0.949) obtained with the same scanner (as training) (p-value: 0.9750).

In contrast, calibration was significantly poorer with the different scanner (Brier score:LJ0.229 [0.124-0.334]) compared to the training-scanner system (Brier score:LJ0.055 [0-0.112]). These findings indicate that *COCOH*’s ability to stratify cancer patient risk is unfazed by scanner variation; however, sites using different scanners may require post-processing calibration to maintain accurate probability estimates.

## Discussion

This multicenter study developed and validated *COCOH*, a multimodal DL framework that integrates disease subtype, histological features, and KRT13-based immunohistochemical features to comprehensively predict cancer risk in OPMD, particularly OL and OLD. To our knowledge, this is the first advanced multimodal system to employ gated attention and two-stage intermediate fusion for combining histopathological and KRT13 immunohistochemical expression features as well as disease subtype, improving accuracy and precision in cancer risk assessment. *COCOH* demonstrated excellent discrimination (AUC: 0.904-0.951), calibration (Brier score: 0.062-0.089), and net clinical benefit for MT prediction in OPMD across testing and prospective validation cohorts. *COCOH*’s performance remained optimal for OL (AUC: 0.897, Brier: 0.113) and OLD (AUC: 0.9, Brier score: 0.047) subtypes of OPMD, with AUPRC values that were higher than random chance. In OL, compared with existing clinical risk assessment methods, *COCOH* had a higher AUC (0.897 vs. 0.753-0.842), recall (85.2% vs 59.3%), and balanced accuracy (85.5 vs. 75.4%) than the WHO and binary dysplasia grading systems. Furthermore, the DL model provided a net reclassification improvement of 20% over binary dysplasia grading for cancer risk stratification in OL. Notably, model discrimination was unaffected by differences in WSI scanners, supporting generalizability across different centers.

Previous studies have employed WSIs for cancer risk assessment in OPMD ^20,28^. Zhang et al^29^ developed an oral mucosa risk stratification (OMRS) model based on patch-level supervision to classify MT risk in OL, where high-risk groups exhibited higher five-year cancer progression rates than low-risk groups. Cai et al ^19^ developed a pathomics-based model that employs convolutional neural networks and conventional machine learning models, achieving an AUC of 0.813 for predicting cancer risk in OL. Shephard et al developed the OMTScore ^17^ and ODYN ^16^ models to predict cancer progression of oral epithelial dysplasia, with AUC values ranging from 0.73 to 0.75. Likewise, Liu-Swetz et al ^18^ developed a model based on multiscale H&E patches encoded using a foundational vision transformer for pathology, achieving an AUC of 0.798. In comparison, *COCOH* significantly outperforms previously described models at comparable validation levels (AUC: 0.904-0.951), likely due to its integration of complementary KRT13 expression data and advanced attention-based pooling framework. Interestingly, *COCOH* does not require manual epithelial annotations or pathologist input to provide risk assessment in OPMD.

Notably, *COCOH* provides simultaneous MT predictions for two OPMD subtypes i.e., OL and OLD by ensuring the implementation of disease information to contextualize training and outputs. Additional strengths of this study encompass *COCOH*’s development using a large sample size from two centers, comprehensive performance analysis including fairness, current standards comparison, alternate architecture evaluations, missing modality assessments, and prospective validation. Explainability was further implemented at multiple modalities, and generalizability was evaluated by site-specific factors analysis.

To facilitate clinical translation, *COCOH* was constructed to primarily provide calibrated MT probabilities for OL and OLD, alongside explainability heatmaps and high-attention patches across multiple H&E scales and WSI modalities These features position the model as a valuable assistive tool facilitating clinical decision-making in OPMD management. However, it should be stated that *COCOH*’s performance may be affected by biopsy procedures, especially when representative areas are not sampled. Our study showed that variations in WSI scanners do not impair model discrimination but could reduce the calibration. This suggests that recalibration of the model or selection of center-specific probability thresholds may be required before clinical implementation in different centers ^30^.

Some limitations exist in this study. While *COCOH* had excellent performance in OL with or without dysplasia, risk stratification is more effective for cases without dysplasia. This discrepancy may reflect the lower proportion of dysplasia cases at training. Future iterations of *COCOH* may mitigate this bias by expanding the dataset to include more OL cases with dysplasia may mitigate this bias. Moreover, while the model included WSIs and cohorts from multiple centers, these were within the same geographic region. Though this study simulated regional differences by varying the WSI scanner to assess *COCOH*’s performance, a broader international evaluation of the model is required to establish generalizability and impact. Last, future updates of *COCOH* could integrate other clinical variables beyond disease subtypes to further refine OPMD cancer risk predictions.

Overall, this study presented *COCOH*, a robust and generalizable multimodal DL system for cancer risk assessment in OPMD (OL and OLD subtypes) that integrates histopathological, immunohistochemical, and disease subtype information. The model provides accurate/well-calibrated risk predictions for MT in OPMD, with no reliance on manual annotations. *COCOH* has strong potential for integration into clinical workflows to streamline prevention strategies and promote earlier detection of oral cancer.

## Acknowledgements

Whole-slide images were scanned at the Imaging and Flow Cytometry Core of the Centre for PanorOmic Sciences (CPOS), Li Ka Shing Faculty of Medicine, University of Hong Kong. The authors are grateful for the technical support provided by Dr Cindy Wang at the facility.

## Data availability

H&E and KRT13 whole-slide images are not publicly available due to restrictions on sharing patients’ sensitive data, in accordance with the provisions of the Institutional Review Board and ethics approval. However, encoded feature vectors (“bags”) may be provided for research purposes only upon reasonable request to the corresponding authors.

## Code availability

Codes to implement *COCOH* have been deposited in the repository available at: https://github.com/HKU-AI4HNO/COCOH.

