## Supplementary Methods for "*COCOH*: A Multimodal Deep Learning Framework for Cancer Risk Assessment of Oral Potentially Malignant Disorders"

<u>**Supplementary Materials**</u>


| **No** | **Contents** | **Page number** |
|---|---|---|
| **1** | **Supplementary Methods** | **2** |
| 1.1 | Data sources | 2 |
| 1.2 | H&E staining and KRT13 immunostaining | 2 |
| 1.3 | Whole-slide imaging | 2 |
| 1.4 | Data preprocessing | 2 |
| 1.5 | *COCOH*’s architecture | 3 |
| 1.6 | Feature encoding | 3 |
| | **Figure 1:** Overview of COCOH’s architecture | 3 |
| 1.7 | Attention-based multiple instance learning | 4 |
| 1.8 | Two-stage multimodal fusion | 4 |
| 1.9 | Model training and cross-validation | 5 |
| | **Table S1**: List of hyperparameters used for tuning *COCOH* during training | 5 |
| 1.10 | Model performance analysis | 5 |
| 1.10.1 | Internal validation and ablation experiments | 5 |
| 1.10.2 | Comparison with alternate architectures | 5 |
| 1.10.3 | Model Testing | 6 |
| 1.10.4 | Benchmarking against dysplasia grading in OL | 6 |
| 1.10.5 | Prospective validation | 6 |
| 1.10.6 | Net benefit and Explainability | 6 |
| 1.10.7 | Site-specific factors analysis | 6 |
| 1.11 | Computation | 7 |
| **2** | **Supplementary results** | **8** |
| 2.1 | Comparison of alternate strategies | 8 |
| | **Table S2**: Count of WSI patches extracted for different datasets | 8 |
| | **Figure S1**: Bar plots showing the performance of different components of *COCOH* with alternative strategies. | 9 |
| | **Figure S2**: Bar plots of *COCOH*’s internal validation performance compared to missing modality and unimodal variants. | 10 |
| | **Figure S3**: Multimodal explainability heatmaps based on *COCOH*’s normalized attention scores for WSIs with misclassifications based on probability thresholds | 11 |
| | **References** | 12 |

### 1. Supplementary methods

#### 1.1 Data sources

This multicenter study utilized a total of 2,656 OL and OLD tissue slides (1,328 stained with H&E and 1,328 underwent KRT13 IHC), sourced from 949 patients across two centers to predict the risk of cancer development in OPMD. The cohort included those with OL and OLD who were managed at the Queen Mary Hospital (QMH) and the Prince Philip Dental Hospital (PPDH) in Hong Kong from January 2003 to December 2022. OLD comprised oral lichen planus and oral lichenoid lesions. The criteria for optimal patient selection were based on the WHO 2020 definition for OPMD [1], and those with other OPMDs or oral malignancies at the time of diagnosis or those without initial biopsies or archival tissues were excluded from this study to obtain the final cohort. For all patients, definitive diagnoses (OL or OLD) and clinical outcomes (cancer development or otherwise as of November 18, 2025) were extracted from the electronic medical records of both institutions. Malignant transformation (MT) was defined as biopsy-proven oral squamous cell carcinoma at the same anatomic site as OPMD after six months of index OPMD biopsy. Those who did not develop cancer had at least 34 months of follow-up. All formalin-fixed paraffin-embedded (FFPE) tissues from surgical biopsies of patients performed before cancer development or the last follow-up were obtained. Characteristics of the study cohort have been detailed in a previous study [2]. Overall, 534 patients with OL (781 tissue samples) and 415 patients with OLD (547 tissue samples) with malignant transformation rates of 12.4% and 9.9% were included (mean follow-up of 97.4 months). Malignant transformation rate for OLD may be elevated in this study due to patient selection according to FFPE availability, which may enrich for lesions of clinical concern. Ethics approval was obtained from the Institutional Review Board of the University of Hong Kong/Hospital Authority Hong Kong West Cluster (Reference no: UW 22-789), and informed consent was waived due to the retrospective nature of this study.

#### 1.2 H&E staining and KRT13 immunostaining

Serial 5μm FFPE sections of OL and OLD from the QMH and PPDH cohorts were processed for H&E staining and immunostaining. H&E staining procedures included deparaffinization with xylene (twice) and graded alcohol (100% (twice), 95%, and 70%), staining with hematoxylin for 3 minutes, differentiation using 0.1% HCl in 70% ethanol for 3 seconds, bluing in 1% ammonia for 1 minute, and additional staining with Eosin Y for 40 seconds. Slides were then air-dried and mounted with coverslips. KRT13 IHC procedures have been documented in a previous report [2]. In summary, after deparaffinization, we performed heat-induced epitope retrieval (HIER) with citrate buffer (0.05% Tween 20, pH 6), peroxidase blocking by incubating in 3% hydrogen peroxide, blocking of non-specific antibody binding using 10% normal goat serum (NGS), overnight 4°C primary antibody incubation (recombinant rabbit monoclonal anti-cytokeratin 13 antibody, clone: EPR3672, 1:100, *Abcam, Cambridge, UK*) incubation, secondary antibody incubation for 1 hour (goat anti-rabbit HRP-conjugated antibody, 1:300, Abcam, Cambridge, UK), and 3,3'-diaminobenzidine (DAB) incubation for visualization. IHC sections were then counterstained with hematoxylin, dehydrated in graded alcohol, cleared in xylene, and mounted with coverslips.

#### 1.3 Whole-slide imaging

H&E (n=1328) and KRT13 IHC (n=1328) sections were digitized using the brightfield settings of the high-throughput PhenoImager HT system (Akoya Biosciences, MA, USA) at 40× magnification with a resolution of 0.25μm/pixel. Other resolutions at 20×, 10×, and 5× were 0.5 μm/pixel, 1 μm/pixel, and 2 μm/pixel. WSIs were in the QPTIFF format (pyramidal), which is a proprietary file containing the scanned pyramidal image with additional metadata. Overall, 2,656 WSIs were scanned and used for *COCOH*'s development and validation.

#### 1.4 Data preprocessing

We performed label encoding for disease information and MT status in spreadsheets. H&E and KRT13 WSIs were anonymized and matched to annotated patient outcomes (malignant transformation) and disease information (OL or OLD) recorded in a spreadsheet. WSIs were loaded using Openslide and we performed Otsu's thresholding to delineate tissue areas and remove background. Nonoverlapping patches of 256 × 256 pixels were then extracted from tissue areas with only those containing <90% white background retained following tile post-processing. Multiscale tiling (5×, 10×, and 20×) was performed for H&E WSIs to ensure context awareness and representation of architectural and cytologic features, while mimicking the workflow of pathologists when reviewing histopathology slides of patients with OPMD [3]. Stain normalization was not performed to mitigate batch- and site-specific factors in model performance [4]. For KRT13 WSIs we only performed single-scale tiling at 5× since the protein is an epithelial differentiation marker that indicate cancer progression with decreasing expression that would be apparent at low-level magnification {Adeoye, 2025 #22;Wils, 2020 #23}. Images were resampled offline using a stratified 75:15:10 train-validation-test split at the patient level to prevent data leakage. Patients were first stratified by the type of OPMD (OL or OLD) and further divided within each strata by their cancer progression status. Afterward, the splitting proportion was applied to ensure that the prevalence of cancer development in OPMD was retained for each disease subtype.

#### 1.5 *COCOH*'s architecture

*COCOH* is a multimodal deep learning framework that integrates disease information (OL or OLD), histopathological image features, and KRT13 IHC image features for cancer risk assessment in OPMD. *COCOH*'s framework uses a hybrid supervised learning approach where OPMD clinical subtypes are implemented using fully supervised learning while histopathological and IHC features are represented using attention-based weakly supervised learning, where the WSIs are viewed as bags of extracted features from H&E/KRT13 IHC image tiles that are aggregated independently and modeled to outcomes that are only available at the WSI level. Furthermore, to enhance cancer risk assessment, the model leverages a multimodal fusion strategy with two modules where histopathological and IHC features are first integrated via intermediate fusion after which the combined WSI representations are fused with the disease identifier before being passed to the classifier head to estimate the probability of malignant transformation of the OPMD. Overview of *COCOH* is presented in **Figure 1**.

#### 1.6 Feature encoding

To generate informative feature representations from H&E and KRT13 IHC image tiles, we used Hibou-B [5], a pathology-specific foundational model based on the Vision Transformer (base variant) architecture and pretrained with the DINOv2 self-supervised learning method on over 1 million diverse WSIs. This model was selected because its pretraining dataset contains both H&E and non-H&E WSIs with a large proportion of WSIs from head and neck tissues compared to other publicly available models. During extraction, each H&E/KRT13 IHC image patches were resized and normalized. Hibou was then run in evaluation mode to convert preprocessed tiles to a feature embedding of 768 dimensions obtained from the [CLS] token of the final transformer layer, which serves as global representation of the WSI patch. This process was then repeated for all the patches from a given WSI ($W$), and the resulting feature vectors were stacked into an $N \times 768$ feature matrix (i.e., bags of features), where $N$ is the total number of image patches obtained from $W$.

**Figure 1:** Overview of COCOH's architecture. The model takes as input the clinical subtype of OPMD and digitized hematoxylin and eosin (H&E) and KRT13 immunohistochemistry (IHC) whole-slide images (WSIs). Disease information is processed through a fully connected layer to generate a disease embedding. H&E WSIs are tiled at multiple resolutions (5×, 10×, and 20×), while KRT13 WSIs are tiled at 5×. Hibou-B serves as the feature encoder, and patch embeddings from each scale or modality are aggregated using attention-based multiple-instance learning. Attention-weighted vectors are concatenated and passed through a fusion module to produce a multimodal WSI-level vector, which is then fused with the disease embedding. The final combined vector is fed into a classifier head to estimate the probability of malignant transformation. Explainability heatmap is generated using patch-level attention scores for each modality.

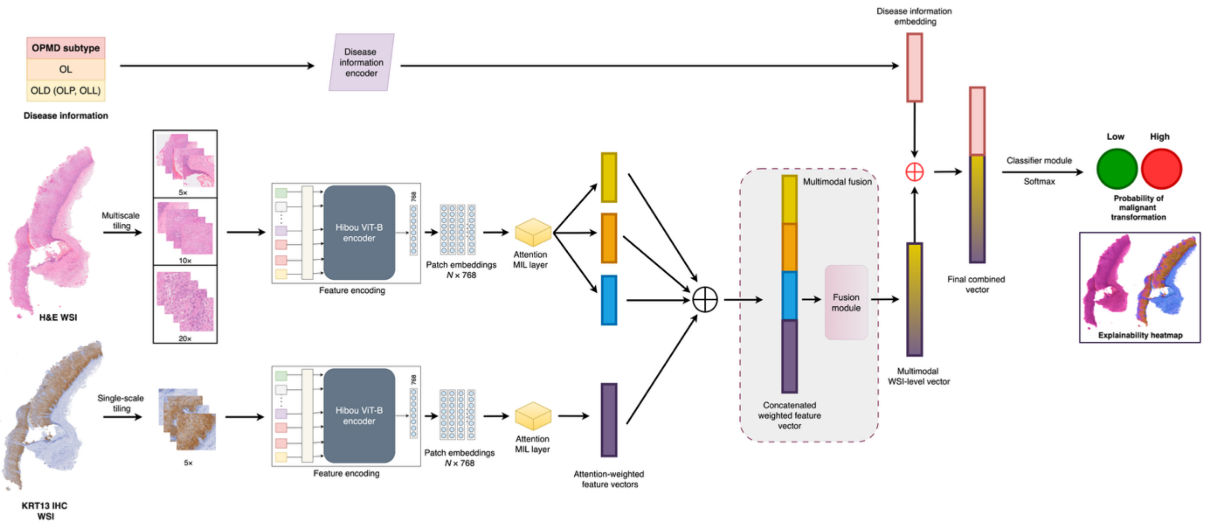

### 1.7 Attention-based multiple instance learning

For H&E and KRT13 WSI modalities, we used tanh-sigmoid gated attention MIL pooling inspired by Ilse et al [6] to aggregate the patch-level features to predict the probability of malignant transformation at the slide level. This trainable deep MIL method uses two parallel neural-network branches with hyperbolic tangent (tanh) and sigmoid activation functions that compute an intermediate attention vector and a gate vector of the same dimension (to modulate feature dimension importance), which are then multiplied elementwise. Gated attention vector is then passed through a final dense layer to compute scalar raw attention scores (unnormalized) for each instance (feature embeddings) in the WSI that are then softmax-normalized and used to weigh and scale patch feature embeddings according to their importance to OPMD outcome prediction. Weighted feature embeddings are summed to obtain WSI-level representations for histopathology and IHC modalities.

Theoretically,

$$\text{Attention scores, } a_i = \frac{\exp(\boldsymbol{w}^{\mathrm{T}}(\tanh(h_i\mathbb{V}) \odot \sigma(h_i\mathbb{U})))}{\sum_{j=1}^{N}\exp(\boldsymbol{w}^{\mathrm{T}}(\tanh(h_j\mathbb{V}) \odot \sigma(h_j\mathbb{U})))}$$

Given:

$N$ = number of instances per bag
$D$ = hidden dimension of instance features
$L$ = hidden dimension of the attention mechanism
$H = [h_1, h_2, h_3, \dots, h_i] \in \mathbb{R}^{N \times D}$ are $N$ instance features (for each WSI) projected to $D$.

Let:

Tangent transformation ($\tau$) for instance ($h_i$) = $\tanh(h_i\mathbb{V}) \in \mathbb{R}^L$, where $\mathbb{V} \in \mathbb{R}^{D\times L}$ is the learnable weight matrix for applying tanh non-linearity.

Sigmoid transformation ($\Sigma$) for $h_i = \sigma(h_i\mathbb{U}) \in \mathbb{R}^L$, where $\mathbb{U} \in \mathbb{R}^{D\times L}$ is the learnable weight matrix for applying sigmoid non-linearity.

Then:

$G_i = \tau_i \odot \Sigma_i$ , where $G_i$ is the gated vector and $\odot$ denotes element-wise multiplication

Raw attention scores, $a_i^r = \boldsymbol{w}^{\mathrm{T}} G_i$, where $\boldsymbol{w} \in \mathbb{R}^{L\times 1}$ is a learnable parameter for computing scalar attention weights for instance.

Normalized attention scores, $a_i = \frac{\exp(a_i^r)}{\sum_{j=1}^{N}\exp(a_j^r)}$

To obtain WSI-level representation, given attention scores $a$ and feature embeddings $H$, MIL aggregation per WSI modality is:

$$z_{modality} = \sum_{i=1}^{N} a_i\, h_i \in \mathbb{R}^D$$

### 1.8 Two-stage multimodal fusion

*COCOH*'s architecture includes a two-stage fusion strategy to integrate the two WSI modalities and disease information (OL or OLD) in this study. First, bag-level representations of WSI modalities ($z_{he}, z_{krt13} \in \mathbb{R}^D$) are concatenated. Given the multiscale aggregated embeddings for the H&E modality (5×, 10×, and 20×), the concatenated WSI-modality representation is implemented as:

$$z_{concat} = \left[z_{he(5\times)};\ z_{he(10\times)};\ z_{he(20\times)}; z_{krt13}\right] \in \mathbb{R}^{\{4D\}}$$

Concatenated representation is then fused via a dense layer with ReLU activation to give a single multimodal WSI-level vector that integrates multiscale H&E and KRT13 modalities which is:

$$z_{wsi} = ReLU\left(z_{concat} \cdot \boldsymbol{w}_{fus}\right) \in \mathbb{R}^D, \text{ where } \boldsymbol{w}_{fus} \in \mathbb{R}^{4D\times D}$$

The scalar disease information ($d \in \mathbb{R}^1$), which is important for contextualizing fused H&E and KRT13 WSI modalities, is passed through a fully connected layer with ReLU activation to output an embedding ($z_{disease}$) projected to $D$.

$$z_{disease} = ReLU(d\ \boldsymbol{w}_{disease}) \in \mathbb{R}^D, \text{ where } \boldsymbol{w}_{disease} \in \mathbb{R}^{1\times D}$$

The multimodal WSI-level vector is then concatenated with the disease information embedding as $z_{combined} = [z_{wsi};\ z_{disease}] \in \mathbb{R}^{2D}$ and this final combined vector ($z_{combined}$) is passed to a classifier head to provide logits that are converted to probabilities of malignant transformation using softmax.

1.9 Model training and cross-validation

To ensure *COCOH*'s stability and select optimal hyperparameters, during training, five-fold cross-validation (CV) was performed only with the training dataset (4 parts training, 1 part validation) while monitoring the loss and area under the precision-recall curve (AUPRC) on the validation partition. List of hyperparameters tuned is in Table S1. Training was achieved with a hidden dimension of 256, batch size of 1, Adam with decoupled weight decay optimizer (AdamW, weight decay=0.00001), maximum epoch of 100, learning rate of 0.0001, dropout of 0.25 and focal loss (modified cross-entropy) to handle imbalance in the outcome data. For the loss function, we set $\alpha$ (balancing) and $\gamma$ (focusing) hyperparameters at 0.75 and 2.0. Early stopping was implemented after a minimum epoch of 5 if no decrease in validation loss is observed after 10 epochs. Mean and standard deviation of the area under the receiver operating characteristic curve (AUC), AUPRC, and Brier scores were calculated across the five folds to determine training performance and stability.

**Table S1**: List of hyperparameters used for tuning *COCOH* during training

| **Hyperparameters** | **Lists** |
|---|---|
| Dimensions ($D$) | 64, 128, 192, 256, 384, 512 |
| Optimizers | Adam, AdamW, SGD |
| Weight decay | 0.001 – 0.000001 |
| Learning rate | 0.01 – 0.00003 |
| Dropout | 0.1 – 0.5 |
| $\alpha$ | 0.5 – 0.9 |
| $\gamma$ | 1 – 3 |

1.10 Model performance analysis

To evaluate COCOH's performance, we retrained the model on the entire training dataset using the same parameters as in CV.

1.10.1 Internal validation and ablation experiments

Discrimination and calibration were assessed on the internal validation dataset using AUC and Brier score. To assess the impact of multimodal WSI fusion to model performance, we conducted ablation experiments by removing different modalities implemented in *COCOH* and determining the AUCs and Brier scores of the resulting model. This involved dropping the modalities by setting $z_{modality}$ to zero in the training and validation datasets. KRT13 WSIs were available for corresponding H&E WSIs per patient, and we did not perform modality dropout during training. However, since KRT13 WSIs may likely be missing compared to H&E in clinical settings, we implemented zero substitution of KRT13 input embeddings with the same shape as the 5× H&E embedding in *COCOH* to allow the model to generate a stable and constant placeholder if this modality is absent. We then computed the AUC and Brier score of the model when all KRT13 WSIs of the validation set were dropped to quantify the impact of this approach on performance at internal validation.

1.10.2 Comparison with alternate architectures

To ensure that the different components and strategies for developing *COCOH* are optimal, this study benchmarked the model performance (AUC and Brier score) against potential architectures trained and evaluated using the internal validation set. Alternative strategies explored include: (i) feature encoders – Virchow [7], CTransPath [8], ResNet50, and EfficientNet-B2 (ii) whole slide-level aggregation - multi-head self-attention and transformer based aggregator (iii) fusion mechanism including early fusion of feature embeddings, hybrid fusion (intermediate fusion of multiscale H&E and weighted late fusion of KRT13 and H&E modalities), and modality-based fusion (learn weights for each modality or resolution using the training data) (iv) Data augmentation techniques including gaussian noise and feature dropout (v) variant model components including GELU instead of ReLU, Adam instead of AdamW, and binary cross-entropy instead of focal loss.

#### 1.10.3 Model Testing

Generalizability assessment was done on the independent testing dataset. Performance metrics for all OPMD and separately for OL and OLD subtypes included AUC, AUPRC, and Brier score. 95% confidence intervals (CI) were also computed for all metrics. Based on Youden's index, we selected optimal thresholds to stratify cancer risk in OL and OLD using the internal validation dataset. This threshold was then applied at testing to calculate the recall, specificity, precision, and negative predictive values of *COCOH*. Differences in AUC and AUPRC between dysplasia status (yes/no) were used to assess potential algorithmic bias in OL predictions.

#### 1.10.4 Benchmarking against dysplasia grading in OL

Since grading dysplasia is the current standard for OL risk assessment, we benchmarked COCOH's test performance against WHO and binary dysplasia grading systems using similar metrics (AUC, recall, specificity, precision, and negative predictive value). Net reclassification improvement (NRI) of *COCOH* was also determined compared to binary grading for patients with OL. Mathematically, NRI is represented as:

$$NRI_{overall} = NRI_{MT} + NRI_{No-MT}$$

Where:

$$NRI_{MT} = \frac{\begin{pmatrix} Number\ of\ cases\ with\ MT\ classified\ as \\ high\ risk\ by\ COCOH \\ and\ low\ risk\ by\ binary\ grading \end{pmatrix} - \begin{pmatrix} Number\ of\ cases\ with\ MT\ classified\ as \\ high\ risk\ by\ binary\ grading \\ and\ low\ risk\ by\ COCOH \end{pmatrix}}{Total\ number\ of\ cases\ with\ MT}$$

$$NRI_{No-MT} = \frac{\begin{pmatrix} Number\ of\ cases\ without\ MT\ classified\ as \\ low\ risk\ by\ COCOH \\ and\ high\ risk\ by\ binary\ grading \end{pmatrix} - \begin{pmatrix} Number\ of\ cases\ without\ MT\ classified\ as \\ low\ risk\ by\ binary\ grading \\ and\ high\ risk\ by\ COCOH \end{pmatrix}}{Total\ number\ of\ cases\ without\ MT}$$

#### 1.10.5 Prospective validation

Additional evaluation of the model was performed using an independent cohort of 41 consecutive patients with OPMD (124 tissue slides, 62 per modality) treated at PPDH between January 1 and December 31, 2023. This cohort had a 4.9% event rate and an average follow-up of 27.3 months. H&E staining, KRT13 IHC, and whole-slide imaging were conducted using protocols similar to those of the development cohort (about 18 months after dataset development for the training, validation, and testing dataset). Using this dataset, we determined the discrimination and calibration performance of *COCOH* using similar metrics above.

#### 1.10.6 Net benefit and Explainability

Net benefit was determined separately for patients with OL and OLD using decision curve analysis of predicted probabilities obtained for patients in the testing and prospective validation dataset. All possible threshold probabilities were explored, and all values were used to plot decision curves for COCOH, which were compared to reference decision curves. Explainability for *COCOH* was implemented at the 5× and 20× scales of the H&E modality and 5× scale of the KRT13 modality using intrinsic attention weights computed for each patch. For each WSI, patch-level attention scores were obtained from the trained model, normalized using percentile scaling, and interpolated to the original resolution to generate a scaled probability heatmap for visualizing patches that contributed to the predicted probabilities. Patch probability heatmaps were then superimposed on WSIs per modality. WSIs of ten random patients (5 OL and 5 OLD, 60% event rate) in the independent testing cohort (whose predictions were correct) were reviewed by two pathologists (AWIL and USK) to identify histologic and immunohistochemical features associated with high or low malignant transformation probability. Likewise, 7 patients (5 OL and 2 OLD, 42.9% event rate) in the same cohort whose predicted probabilities were incorrect were reviewed to identify histologic and immunohistochemical features in the WSIs contributing to misclassification. To confirm importance of patch-level features, we removed the vectors with the top ten attention scores per modality for the test and prospective validation data before recalculating the AUC and Brier scores to ensure these areas accurately informed predictive performance.

##### 1.10.7 Site-specific factors analysis

To evaluate the impact of whole-slide imaging systems on model performance, we randomly selected 124 tissue slides (62 H&E and 62 KRT13) from the test and prospective validation sets and rescanned them at 20× on a Nikon Eclipse Ti2-E inverted microscope. WSIs (TIFF) were tiled into 256×256 patches. Also, WSIs were downsampled by factors of two and four during tiling to yield 10× and 5× patches. Patches were encoded using Hibou-B and processed via *COCOH*'s attention MIL pooling, modality fusion, and prediction module. Performance on these rescanned WSIs were assessed using AUC and Brier score and compared with WSIs of the same slides (matched by slide ID) acquired with the Akoya PhenoImager HT system. Differences in the AUC of the model between the imaging modalities were assessed using DeLong's test, with p-values below 0.05 considered as statistically significant. 95% confidence intervals were also determined for the differences in AUC and Brier score.

#### 1.11 Computation

Data processing and model development were conducted using Python 3.11 libraries including OpenSlide, OpenCV, PIL, scikit-image, and PyTorch. Training, validation, and explainability were achieved using a workstation with an Nvidia RTX A4500 GPU. Decision curve analysis and threshold selection were performed in R v 4.1.2 using the dcurves and cutpointr packages.

### 2.0 Supplementary results

**Table S2**: Count of WSI patches extracted for different datasets

| Dataset | **5× H&E** | **10× H&E** | **20× H&E** | **KRT13** | **Total** |
|---|---|---|---|---|---|
| Training | 101,645 | 362,245 | 1,358,624 | 104,438 | 1,926,952 |
| Validation (internal) | 23,912 | 86,816 | 330,062 | 24,331 | 465,121 |
| Test | 12,367 | 44,507 | 166,280 | 12,928 | 236,082 |
| Validation (prospective) | 6,039 | 21,506 | 82,469 | 6,125 | 116,139 |
| Total | 143,963 | 515,074 | 1,937,435 | 147,822 | **2,744,294** |

#### 2.1 Comparison of alternate strategies

Alternative approaches were compared to ensure that the different components of *COCOH* are optimal. None of the comparator feature encoders outperformed Hibou-B, with Virchow being the best alternative for discrimination (AUC: 0.902) and ResNet 50 the best alternative for calibration (Brier score: 0.078) (**Fig. S1A, B**). The tanh-sigmoid attention mechanism performed similarly to complex techniques like multi-head self-attention and transformer-based aggregation (**Fig. S1C, D**). The two-stage fusion method employed in *COCOH* also yielded the best AUC and Brier score compared to modality-based fusion (AUC: 0.908, Brier score: 0.077) and hybrid fusion (AUC: 0.9, Brier score: 0.076) (**Fig. S1E, F**). Data augmentation techniques did not improve model discrimination and even worsened calibration, which does not justify the added complexity (**Fig. S1G, H**). Also, tuning the activation function and optimizer did not change the AUC or Brier score of the model; however, when binary crossentropy was set as the loss function, the model's AUC decreased by 0.002 while the Brier score increased by 0.007 (**Fig. S2I, J**).

**Figure S1**: Bar plots showing the performance of different components of *COCOH* with alternative strategies. (A) **AUC** of Hibou-B compared to other pathology-specific and conventional **feature encoders** (B) **Brier scores** of Hibou-B compared to other pathology-specific and conventional **feature encoders** (C) **AUC** of *COCOH* when gated attention mechanism was used compared to other **weakly-supervised methods** (D) **Brier score** of *COCOH* when gated attention mechanism was used compared to other **weakly-supervised methods** (E) **AUC** of *COCOH* when two-stage fusion was used to integrate modalities compared to other **fusion techniques** (F) **Brier score** of *COCOH* when two-stage fusion was used to integrate modalities compared to other **fusion techniques** (G) **AUC** of *COCOH* when training dataset was **augmented vs no augmentation** (H) **Brier score** of *COCOH* when training dataset was **augmented vs no augmentation** (I) **AUC** of *COCOH* when different **hyperparameters** were changed (J) **Brier score** of *COCOH* when different **hyperparameters** were changed.

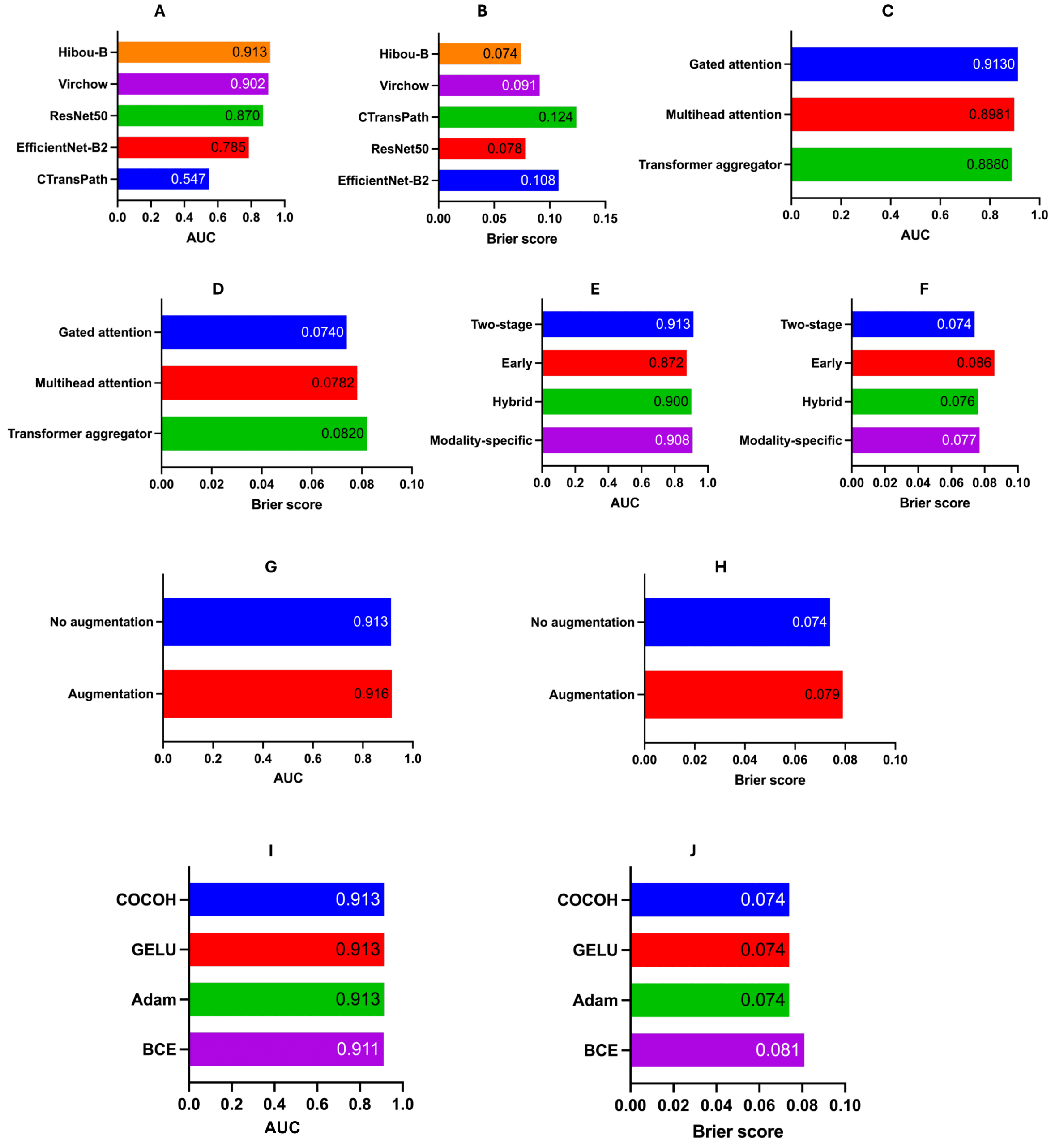

**Figure S2**: Bar plots of *COCOH*'s internal validation performance compared to missing modality and unimodal variants. (A) **AUC** of the original model and variant models based on zero KRT13 vectors and H&E features alone for predicting cancer risk in **OPMD**. (B) **AUPRC** of the original model and variant models based on zero KRT13 vectors and H&E features alone for predicting cancer risk in **OPMD**. (C) **Brier score** of the original model and variant models based on zero KRT13 vectors and H&E features alone for predicting cancer risk in **OPMD**. (D) **AUC** of the original model and variant models based on zero KRT13 vectors and H&E features alone for predicting cancer risk in **OL**. (E) **AUPRC** of the original model and variant models based on zero KRT13 vectors and H&E features alone for predicting cancer risk in **OL**. (F) **Brier score** of the original model and variant models based on zero KRT13 vectors and H&E features alone for predicting cancer risk in **OL**. (G) **AUC** of the original model and variant models based on zero KRT13 vectors and H&E features alone for predicting cancer risk in **OLD**. (H) **AUPRC** of the original model and variant models based on zero KRT13 vectors and H&E features alone for predicting cancer risk in **OLD**. (I) **Brier score** of the original model and variant models based on zero KRT13 vectors and H&E features alone for predicting cancer risk in **OLD**.

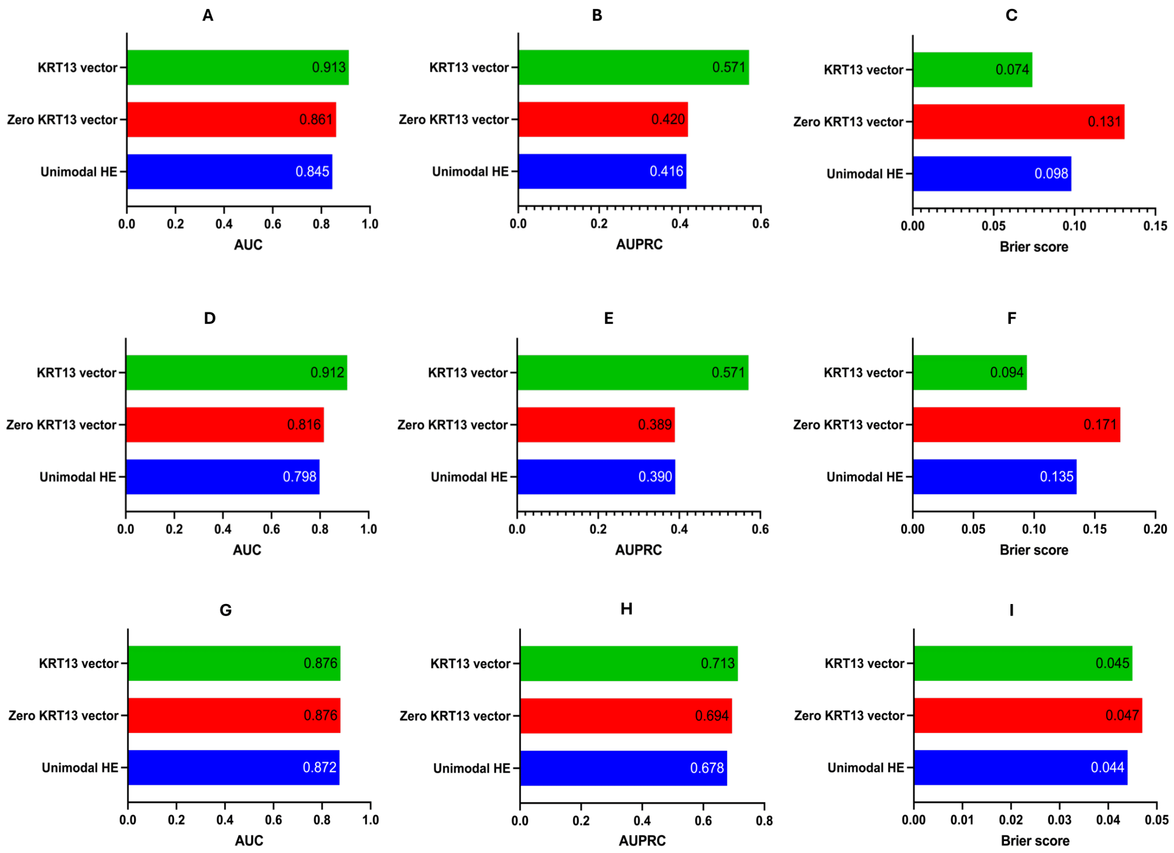

**Figure S3**: Multimodal explainability heatmaps based on *COCOH*'s normalized attention scores for WSIs with misclassifications based on probability thresholds. (A) H&E WSI for an OLD case without MT after 32 months follow-up. *COCOH* predicted an MT probability of 61% (B) Corresponding KRT13 IHC WSI of patient in A. (C) Attention heatmaps of H&E WSIs at 5× resolution (D) Attention heatmaps of H&E WSIs at 20× resolution (E) Attention heatmaps of KRT13 WSIs at 5× resolution (F) H&E WSI for an OL case with MT after 18 months follow-up. *COCOH* predicted an MT probability of 4.8% (G) Corresponding KRT13 IHC WSI of patient in F (H) Attention heatmaps of H&E WSIs at 5× resolution (I) Attention heatmaps of H&E WSIs at 20× resolution (J) Attention heatmaps of KRT13 WSIs at 5× resolution.

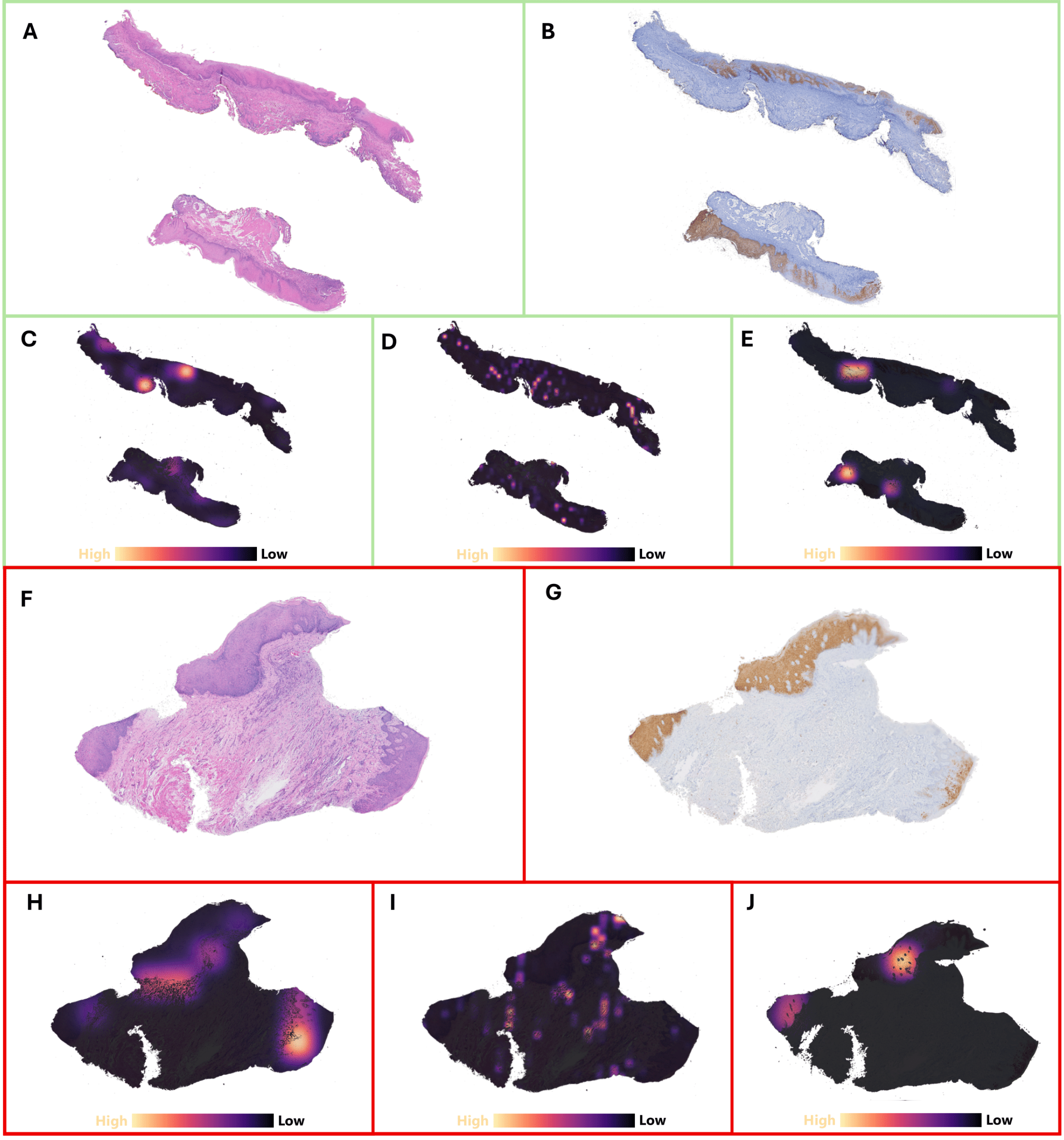
